# Back to school: use of Dried Blood Spot for the detection of SARS-CoV-2-specific immunoglobulin G (IgG) among schoolchildren in Milan, Italy

**DOI:** 10.1101/2020.07.29.20164186

**Authors:** Antonella Amendola, Silvia Bianchi, Maria Gori, Lucia Barcellini, Daniela Colzani, Marta Canuti, Vania Giacomet, Valentina Fabiano, Laura Folgori, Gian Vincenzo Zuccotti, Elisabetta Tanzi

**Author notes:** **Corresponding author**: Prof. Antonella Amendola, Department of Biomedical Sciences for Health, University of Milan, Milan, Italy, 0039 02 503 25 072.

## Abstract

Serological surveillance is necessary to the reestablishment of school activities in safe conditions and to avoid school-related outbreaks. In this study, DBS (Dried Blood Spots) have proven to be a simple, rapid and reliable sample collection tool for detecting antibodies against SARS-CoV-2 by ELISA test compared to matched serum samples from venous sampling (R2=0.9553; Pearson’s coefficient=0.98; Cohen’s unweighted k=0.93; overall agreement=96.2%). This approach may facilitate sample collection from schoolchildren for serological surveys useful to an adequate risk-assessment.

## Introduction

The pandemic spread of coronavirus disease 2019 (COVID-19) has resulted in the temporary closure of schools in most countries, affecting approximately 1.6 billion children and young people and 91% of students worldwide. These unprecedented numbers and the enormous implications following school closures immediately fuelled a heated debate [1].

International data on the spread of COVID-19 seems to indicate that younger individuals are less susceptible to the disease. However, only a little is known about the real impact of SARS-CoV-2 infection in the paediatric population and it is not yet clear whether children become less infected than adults and/or are less able to spread the virus [2].

In this scenario, serological studies play the crucial role of facilitating adequate public health interventions and the reorganization of school activities in safe conditions [3]. The most used serological assays, enzyme-linked immunosorbent assays (ELISA) and chemoluminescence assays (CLIA) [4], detect antibodies against SARS-CoV-2 in sera from venous blood collection, an invasive sample collection method that may be of difficult implementation in a paediatric setting. Therefore, to be able to test the largest possible number of children and adolescents, the availability of a minimally invasive sampling method is fundamental. Rapid tests on capillary blood could be a convenient alternative, however they are not always accurate. Indeed, several Health Authorities have warned against the use of rapid tests, requiring confirmation of positivity by comparison with a gold standard test, or even prohibited them [4]. The use of Dried Blood Spots (DBS), drops of capillary blood dried on filter paper, allows to overcome problems connected to sample collection and, at the same time, facilitates sample transport and storage [5-7]. DBS are frequently used in clinical testing and for surveillance studies and in 2007 the World Health Organization (WHO) published recommendations for the use of DBS as sample collection method for detecting protective antibodies against measles and rubella [8]. In this study, we assessed the adequacy of DBS as a tool to collect samples for the detection of SARS-CoV-2 type G immunoglobulins (IgG) via a commercially available (CE approved) semi-quantitative ELISA.

## Materials and Methods

The performance of DBS as a means of collecting samples to measure SARS-CoV-2 specific IgG was tested by comparing serologic results obtained from matched serum and DBS samples from 52 Health Care Workers (HCWs). HCWs were asymptomatic at the time of sample collection and all worked in a tertiary children hospital in Milan. Ethics approval was obtained from the Ethics Committee of the Buzzi Hospital (Prot. N 0018927); data were collected, and samples handled according to the Declaration of Helsinki, as revised in 2013.

Drops of capillary blood from a finger prick were collected (two saturated spots on one paper) on cellulose-based DBS cards (Perkin-Elmer 226 filter paper, Greenville, SC, USA). DBS cards were air-dried for a couple of hours and successively stored in plastic bags (with desiccant to reduce humidity) at room temperature. Four 3 mm disks were punched out and incubated for 1 hour at room temperature in 500 µL of ELISA sample buffer with constant shaking (400 rpm) to elute antibodies from the paper. DBS eluates and paired sera (100 µl of serum diluted 1:101 in sample buffer) were tested simultaneously for the presence of anti-SARS-CoV-2 IgG with a commercially available ELISA (Euroimmun Medizinische Labordiagnostika, Lubeck, Germany) according to the manufacturer’s instructions. Samples were classified according to ratio values of sample absorbance over calibrator (ODs/Cal) as recommended by the manufacturer into three categories: negative (<0.8), inconclusive (≥0.8 and <1.1), and positive (≥1.1). The Pearson’s coefficient was calculated to evaluate concordance between measures and results were considered statistically significant at p<0.05. Concordance between tests was assessed using the Kappa statistic (Cohen’s unweighted Kappa, k) as: poor (k=0), slight (0.01<k<0.20), fair (0.21<k<0.40), moderate (0.41<k<0.60), substantial (0.61<k<0.80), almost perfect (0.81<k<1.00), or perfect (k=1). Linear modelling for regression analysis was performed and visualized in R [9, 10].

## Results

Twenty-three serum samples resulted IgG-positive (ODs/Cal: 1.1-6.2), 28 were negative (ODs/Cal: 0.1-0.7 ODs/Cal), and one test was inconclusive (0.9 ODs/Cal). Similarly, 21 DBS samples were considered positive (ODs/Cal:1.1-6.1), 28 negative (ODs/Cal: 0.2-0.7), and 3 tests were inconclusive (ODs/Cal: 0.8-1.0). Overall, only 2 discordant results were obtained and in both cases serum samples tested positive, while the tests performed on DBS eluates were inconclusive. Results are summarized in Table 1.

**Table 1.**
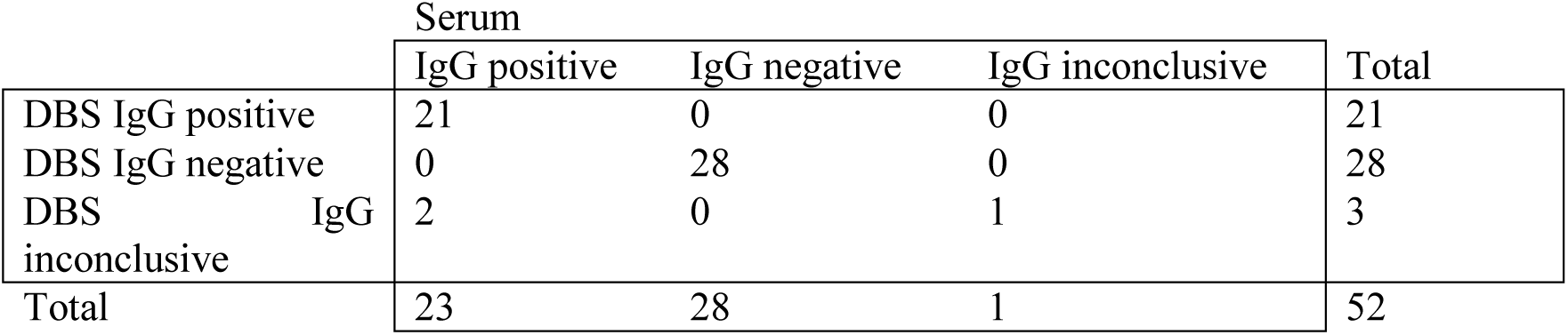
Summary of anti-SARS-CoV-2 IgG results among paired sera and DBS eluates.

|  | Serum |  |  | Total |
| --- | --- | --- | --- | --- |
|  | IgG positive | IgG negative | IgG inconclusive |  |
| DBS IgG positive | 21 | 0 | 0 | 21 |
| DBS IgG negative | 0 | 28 | 0 | 28 |
| DBS IgG inconclusive | 2 | 0 | 1 | 3 |
| Total | 23 | 28 | 1 | 52 |

The correlation coefficient (R^2^) between the two sets of samples was 0.9553, the Pearson’s coefficient was 0,98 (0.97-1, p<0.0001), k was 0.93 (0.92-1, p<0.0001), and the overall agreement was 96.2% (86.8-99.5). The linear relation analysis between the ODs/Cal values obtained from matched DBS and serum samples is shown in Figure 1.

**Figure 1.**
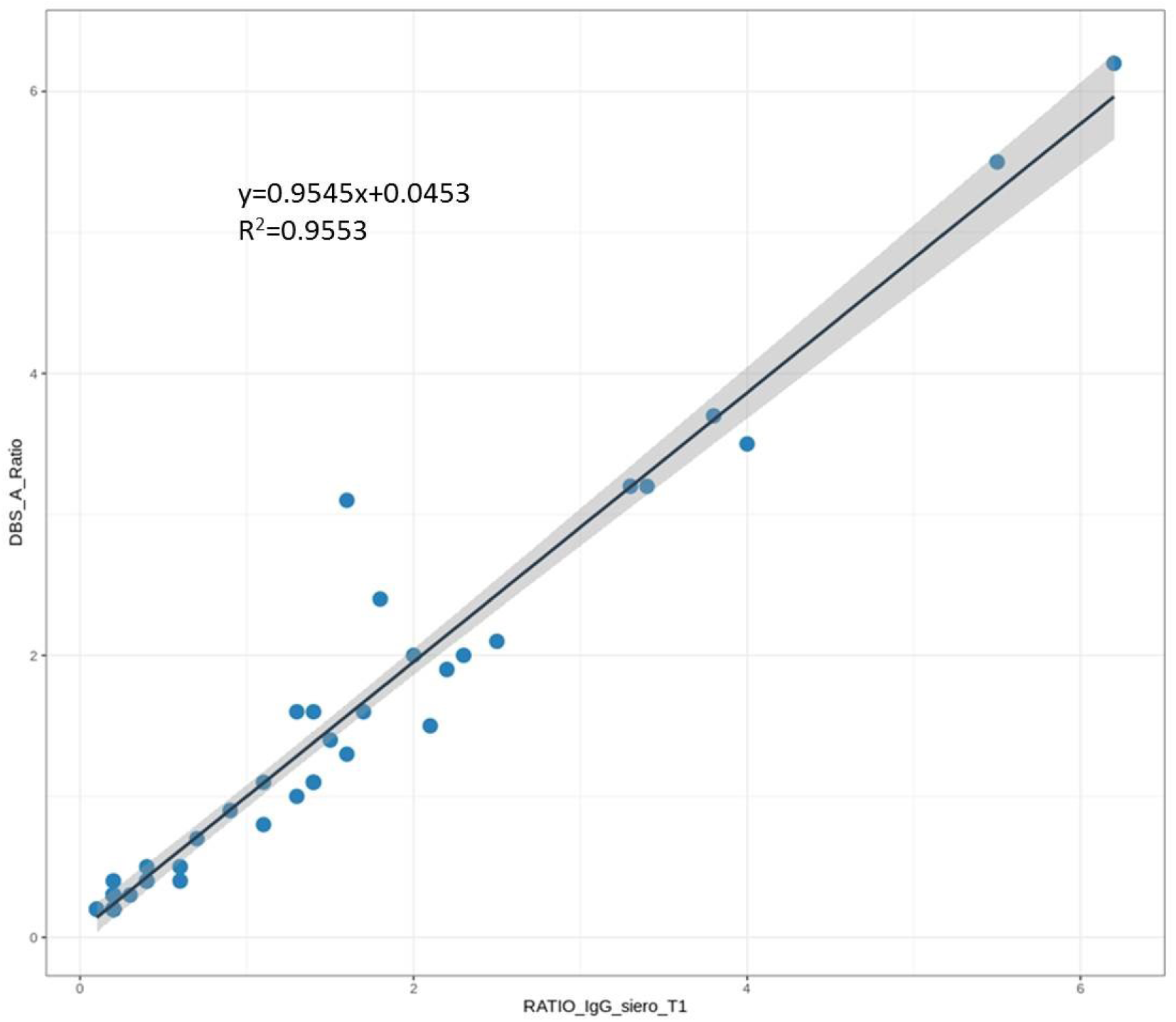
The linear regression between results obtained with sera and DBS eluates for 52 paired samples. Also shown are observations (dots) and 95% confidence interval (grey).

## Conclusions

To the best of our knowledge, this is the first report of anti-SARS-CoV-2 IgG detection from dried blood spots stored on filter paper compared with serum. The analysis of both positive and negative samples demonstrated the validity of DBS as sampling methods for anti-SARS-CoV-2 IgG detection by a commercial ELISA. The data acquired showed an elevated concordance between results obtained with DBS eluates and serum samples and Cohen’s unweighted Kappa values were very high, indicating an “almost perfect” agreement. These data support the feasibility of the use of DBS as a simple, rapid, and reliable sample collection tool for the detection of antibodies against SARS-CoV-2 [11].

DBS represent a minimally invasive blood sampling procedure that combines the convenience of sample collection in non-clinical settings [5-7] with the accuracy of the ELISA in the laboratory. Because of the minimal invasiveness, this approach could facilitate sample collection from schoolchildren for serological surveys and we are planning on using this approach for monitoring the spread of SARS-CoV-2 among children upon their return to schools in Milan, one of the COVID-19 most affected areas during the beginning of the pandemic. The consequent prompt knowledge of the local epidemiological profile will facilitate the reestablishment of school activities in safe conditions, avoiding school-related outbreaks, and the development of targeted solutions aimed at contrasting and containing the negative effects of any epidemic ups and downs. A high-quality surveillance system is essential for an adequate risk-assessment and for developing targeted responses to health emergencies that are calibrated to territorial needs. For this purpose, the availability of a non-invasive sampling method that increases participant engagement is crucial. This will minimize the risk of measures that require drastic responses, such as further prolonged school closures.

## Data Availability

All relevant data are within the manuscript.

## References

[1] UNICEF. Coronavirus disease (COVID-19) information centre. [Accessed on 06 July 2020] Available from: https://www.unicef.org/coronavirus/.

[2] Mallapaty S. How do children spread the coronavirus? The science still isn’t clear. Nature. 2020;581(7807):127–128. doi:10.1038/d41586-020-01354-0.

[3] Caini S, Bellerba F, Corso F, Díaz-Basabe A, Natoli G, Paget J, Facciotti F, De Angelis SP, Raimondi S, Palli D, Mazzarella L, Pelicci PG, Vineis P, Gandini S. Meta-analysis of diagnostic performance of serological tests for SARS-CoV-2 antibodies up to 25 April 2020 and public health implications. Euro Surveill. 2020;25(23):ppii=2000980. https://doi.org/10.2807/1560-7917.

[4] European Centre for Disease Prevention and Control. An overview of the rapid test situation for COVID-19 diagnosis in the EU/EEA.1April2020.Stockholm: ECDC; 2020.

[5] Centers for Disease Control and Prevention. Shipping guidelines for dried-blood spot specimens. [Accessed on 16 June 2020] Available from: https://www.cdc.gov/labstandards/pdf/nsqap/Bloodspot_Transportation_Guidelines.pdf. Published 2017.

[6] Su X, Carlson BF, Wang X, Li X, Zhang Y, Montgomery JP, Ding Y, Wagner AL, Gillespie B, Boulton ML. Dried blood spots: An evaluation of utility in the field. J Infect Public Health. 2018;11(3):373–376. doi:10.1016/j.jiph.2017.09.014

[7] García-Cisneros S, Sánchez-Alemán MÁ, Conde-Glez CJ, Lara-Zaragoza SJ, Herrera-Ortiz A, Plett-Torres T, Olamendi-Portugal M. Performance of ELISA and Western blot to detect antibodies against HSV-2 using dried blood spots. J Infect Public Health. 2019;12(2):224–228. doi:10.1016/j.jiph.2018.10.007

[8] World Health Organization. Manual for the laboratory diagnosis of measles and rubella virus infection, 2nd ed, 2007. [Accessed on 07 June 2020]. Available from: https://apps.who.int/iris/handle/10665/70211.

[9] R Core Team. R: A language and environment for statistical computing. R Foundation for Statistical Computing, Vienna, Austria. 2019. Available from: https://www.R-project.org/.

[10] Wickham H. ggplot2: Elegant Graphics for Data Analysis. Springer-Verlag New York, 2016.

[11] Thevis M, Knoop A, Schaefer MS, Dufaux B, Schrader Y, Thomas A, Geyer H. Can dried blood spots (DBS) contribute to conducting comprehensive SARS-CoV-2 antibody tests?. Drug Test Anal. 2020;12(7):994–997. doi:10.1002/dta.2816Su.

